# Immunogenicity of co-administered vaccine antigens with whole-cell or acellular pertussis vaccines in infancy: a randomised controlled trial

**DOI:** 10.1101/2025.02.16.25321528

**Authors:** Gladymar Pérez Chacón, Sonia McAlister, James Totterdell, Marie J. Estcourt, Julie A. Marsh, Mark Jones, Kirsten P. Perrett, Dianne E. Campbell, Nicholas Wood, Michael Gold, Claire S. Waddington, Michael O’Sullivan, Nigel Curtis, Ushma Wadia, Peter B. McIntyre, Patrick G. Holt, Tom Snelling, Peter C. Richmond

## Abstract

**Background:** Whole-cell pertussis (wP) and acellular pertussis (aP) vaccines evoke different immune responses to pertussis vaccine antigens. We compared the effect of a heterologous wP/aP/aP primary series (hereafter mixed wP/aP) versus a homologous aP/aP/aP primary schedule (hereafter aP-only) on antibody responses to co-administered vaccine antigens in infants and toddlers.

**Methods:** We randomised Australian infants in a 1:1 ratio to receive either a mixed wP/aP schedule (pentavalent diphtheria-tetanus-wP-hepatitis B-*Haemophilus influenzae* type b; DTwP-HepB-Hib vaccine at 6 weeks old followed by hexavalent DTaP-inactivated poliovirus vaccine (IPV)-HepB-Hib vaccine at 4 and 6 months old) or to aP-only priming doses of hexavalent DTaP-IPV-HepB-Hib vaccine at the same ages. All infants received 13-valent pneumococcal conjugate vaccine (13vPCV) at 6 weeks, 4 and 12 months of age and DTaP-IPV and Hib vaccine boosters at 18 months. We estimated the ratio (GMR) of IgG geometric mean concentrations (GMCs) in the wP/aP and aP-only groups for the serotypes included in the 13vPCV, for Hib capsular polysaccharide polyribosylribitol phosphate (PRP), and for hepatitis B surface antigen (HBsAg) at 6, 7, 18, and 19 months. We assessed whether the wP/aP schedule is non-inferior to the aP-only schedule for co-administered vaccine antigens (GMR>2/3). Trial registration: ACTRN12617000065392p.

**Results:** Between March 2018 and January 2020, 150 infants were randomised (75 per study arm). Responses to all 13vPCV serotypes and Hib-PRP at 6, 7, 18, and 19 months old, as well as HBsAg at 6 and 7 months old were non-inferior (>90% probability). Sera GMCs were higher for each 13vPCV serotype, Hib-PRP, and HBsAg at each timepoint in the wP/aP group than in the aP-only group.

**Interpretation:** A mixed wP/aP schedule resulted in non-inferior IgG responses to co-administered vaccine antigens compared to the standard aP-only schedule for pertussis primary immunisation.

**Funding:** Telethon New Children’s Hospital Research Fund and National Health and Medical Research Council.

**Research in context:** *Evidence before this study:* Combination vaccines incorporate antigens that protect against multiple diseases into a single injection. Most low- and middle-income countries (LMICs) currently use wP combination vaccines. Due to the need for periodic boosters to protect older children, adolescents, and adults, these countries may consider moving to the less reactogenic aP combination vaccines that are routinely used in most high-income countries. We searched for evidence about whether a mixed wP/aP primary schedule impacts the immunogenicity of co-administered vaccines. We were particularly interested in evidence for impacts on 13vPCV 2 + 1 schedule and other pneumococcal dose-sparing schedules. We searched PubMed on May 23, 2024, for randomised controlled trials using the following search terms “pneumococcal”, “routine vaccin*”, and “pertussis” combined with Boolean operators, without date or language restrictions. We failed to identify any head-to-head randomised comparisons of the effect of heterologous (mixed) versus homologous pertussis vaccine primary series on co-administered vaccine antigens. Our previous meta-analysis reviewed 15 randomised controlled studies that compared serious adverse events among infants receiving wP versus aP as a first dose before 6 months of age. Few studies reported immune responses to non-DTP co-administered antigens. These findings suggest enhanced Hib responses among recipients of a three-dose primary series of wP compared to those who received three primary aP doses, non-inferior Hib-PRP seroprotection among aP compared to wP vaccinees, and mixed results regarding HBsAg-IgG levels post-wP priming. Both wP and aP groups exhibited weaker Hib-PRP IgG responses when DTP-Hib vaccines were co-administered with meningococcal serogroup C vaccine conjugated to cross-reactive material 197 (CRM_197_) compared to the meningococcal serogroup C vaccine conjugated to tetanus toxoid (TT).

*Added value of this study:* This paper is the first reported evidence of a mixed wP/aP schedule resulting in non-inferior IgG responses to co-administered vaccine antigens compared to the standard homologous aP-only schedule for pertussis primary immunisation. In addition, enhanced immune responses were observed to all serotypes included in the 13vPCV and Hib-PRP vaccines in children receiving the mixed wP/aP vaccination strategy versus those vaccinated with a standard aP-only schedule.

*Implications of all the available evidence:* In settings transitioning from using wP to aP multi-component vaccines, infants receiving a mixed schedule (with wP as the first dose) can be expected have non-inferior, and possibly superior, antibody responses to concomitant vaccine antigens. To better understand the underlying mechanisms of our findings, the assessment of opsonophagocytic activity response rates and serotype-specific memory B cell immune responses to PCV antigens is required. Large population-based studies, particularly in countries where pneumococcal and Hib disease burdens remain high, should be conducted to determine if the observed effects on immune responses translate into differences in protection against disease.

## Introduction

Vaccinating infants with whole-cell pertussis (wP) rather than acellular pertussis (aP) vaccine formulations elicits different, long-lived immune responses and provides enhanced protection against pertussis infection.^1,2^ In recipients of homologous pertussis vaccine priming, the type of pertussis vaccine used as the first dose of the primary series –formulations of diphtheria and tetanus toxoids with either wP (DTwP) or aP (DTaP) – appears to influence T-cell responses to tetanus toxoid (TT) and to diphtheria toxoid (DT).^3,4^

Bacterial capsular polysaccharides, like those in *Streptococcus pneumoniae* or *Haemophilus influenzae* type b (Hib), trigger T-cell independent B-cell responses. In conjugated pneumococcal or Hib vaccines, these polysaccharides are bound to an immunogenic protein carrier (such as TT or CRM_197_, the non-toxic mutant of diphtheria toxin). This binding recruits carrier-specific CD4+ T follicular helper cells, leading to a T-dependent antibody response which is crucial for preventing invasive pneumococcal or Hib disease in infants.^5^ However, immunisation with pertussis vaccines and conjugated polysaccharide vaccines may interact unpredictably.^6^ Interactions are influenced by the type and concentration of protein carriers,^7^ timing and order of vaccination,^8^ coadministration with inactivated poliovirus vaccine (IPV),^9^ off-target responses elicited by *Bordetella pertussis* lipopolysaccharide (present in wP but not in aP vaccines),^10^ and interference from the presence of antibodies prior to vaccination.^11,12^ In contrast, while historical data provide no evidence of a difference in seroprotective responses to simultaneously administered hepatitis B vaccine among recipients of wP versus aP-based primary series,^13^ IgG geometric mean concentrations (GMCs) of the hepatitis B (HepB) surface antigen (HBsAg) appear to be lower among recipients of aP versus wP combination vaccines.^14^

In response to reactogenicity concerns, Australia transitioned from a wP-only to an aP-only primary series in the late 1990s. Observational studies suggest that, among the birth cohorts born during the switchover period from wP-only to aP-only regimens, a first dose of wP, compared to aP, appeared to be protective against the development of childhood IgE-mediated food allergy.^15,16^ The latter remains a major public health problem in this country, affecting up to 10% of infants by age 12 months. Before a novel heterologous pertussis primary vaccination schedule can be recommended, it is important to ensure no unintended adverse impact on the immunogenicity of other routine vaccines, such as pneumococcal conjugate and Hib, and hepatitis B vaccines. We previously reported the non-inferiority of a mixed wP/aP primary schedule compared to the standard aP-only primary schedule with respect to anti-diphtheria, tetanus, and pertussis IgG responses at 7 months old, approximately one month after completion of the primary series.^17^ Here we report and compare antibody responses to concurrently administered 13-valent pneumococcal conjugated vaccine (13vPCV), Hib capsular polysaccharide (Hib-PRP) and HBsAg among infants randomly assigned to either a heterologous wP/aP/aP (hereafter mixed wP/aP) or homologous aP/aP/aP (hereafter aP-only) priming schedule.

## Methods

### Study design and participants

OPTIMUM (Optimising Immunisation Using Mixed Schedules) is a two-stage, Bayesian adaptive, group sequential, randomised, parallel-group, observer-blinded controlled trial.^18,19^ Stage one was conducted in Perth (Western Australia, WA) and planned to compare anti-TT-IgE responses at 7 months old in a cohort of infants randomly assigned to either a mixed wP/aP primary schedule or a standard aP-only primary schedule. In addition, the first stage of this trial compared the reactogenicity and immunogenicity of both schedules.^17^ Stage two will evaluate atopic outcomes (i.e., IgE-mediated food allergy, atopic dermatitis, and atopic sensitisation) by 12 months old in up to 3,000 infants.

In brief, eligible infants were born after 32 weeks’ gestation and aged between 6 and < 12 weeks; full inclusion and exclusion criteria are detailed in the study protocol and statistical analysis plan.^18,19^ Parental consent was obtained immediately before enrolment. Approval was granted by the Child and Adolescent Health Service Ethics Committee, Western Australia (WA), Australia (RGS00019).

### Randomisation and blinding

At 6 weeks old, infants were randomly assigned in a 1:1 ratio to receive either an intramuscular (IM) dose (0·5 mL) of a World Health Organization (WHO)-prequalified pentavalent wP combination vaccine with no IPV components (DTwP-Hib-HepB, Pentabio, PT Bio Farma, Indonesia, hereafter referred to as wP) or the standard hexavalent aP combination vaccine (DTaP-Hib-HepB-IPV, Infanrix Hexa, GlaxoSmithKline, Australia) in the anterolateral aspect of the right thigh.^20^ In both vaccine formulations, Hib-PRP is conjugated to the TT protein.

Randomisation was performed using a computer-generated allocation sequence prepared by the trial statistician, based on randomly permuted blocks of varying size. Details of the allocation concealment were previously reported in the published protocol.^18^

### Procedures

At 4 and 6 months old, all study infants received a dose of the hexavalent aP vaccine per the Australian routine immunisation schedule.^21^ A booster dose of aP was given at 18 months old (0·5 mL; IM) using a DTaP-IPV formulation (Infanrix IPV, GlaxoSmithKline, Australia), thereby preserving blinding while ensuring every participant received a minimum three (and up to four) doses of IPV by 18 months old. All infants were administered two IM priming doses (each 0·5 mL) of the 13vPCV CRM_197_-conjugated vaccine (Pfizer) at 6 weeks and 4 months old, and a booster dose at 12 months old (hereafter referred to as a ‘2 + 1’ schedule). The 13vPCV was injected in the anterolateral aspect of the opposite thigh to co-administered wP or aP vaccines. Additional routine vaccines were administered per national guidelines: oral monovalent rotavirus vaccine at 6 weeks and 4 months old; measles-mumps-rubella combination vaccine and quadrivalent meningococcal (serogroups A, C, W, Y)–TT conjugate vaccine at 12 months old; Hib-PRP conjugated to TT and measles-mumps-rubella-varicella at 18 months old. As a study specific procedure and in alignment with Australian immunisation guidelines from the wP vaccine era, oral paracetamol (15 mg/Kg) was given just before the 6-week vaccine doses to attenuate wP reactogenicity; caregivers were advised to administer two further unobserved doses of paracetamol 6 hours apart. The sampling schedule was aligned with the schedule for pertussis vaccine doses; bloods were collected immediately before the 6-month dose of aP and 4 weeks afterwards (i.e. 8 and 12 weeks after the 4-month dose of the 13vPCV; up to 5 mL), and immediately before the booster dose of DTaP-IPV and Hib vaccines at age 18 months and 4 weeks afterwards (i.e. 6 and 7 months after the 12-month booster dose of the 13vPCV; up to 10 mL).

Serum serotype-specific IgG to pneumococcal capsular polysaccharides included in the 13vPCV (1, 3, 4, 5, 6A, 6B, 7F, 9V, 14, 18C, 19A, 19F, and 23F) were measured at 6, 7, 18, and 19 months old using an inhouse multiplex fluorescent bead-based immunoassay, as previously described, with modifications.^22–25^ In brief, Bio-Plex® COOH-microspheres (6.25 x 10^6^, 500 µl, Bio-Rad) were conjugated overnight with optimised doses of DMTMM-modified pneumococcal polysaccharides (ATCC, Manassas, USA) in 1X PBS pH 7.2 (Life Technologies, Carlsbad, USA) as per Schlottmann et al.^26^ The optimal coating doses were: a 1:1 dose; 500 µl microspheres with 500 µl DMTMM-modified polysaccharides for serotypes 1, 3, 5, 7F and 9V; and a 1:0.5 dose; 500 µl microspheres with 250 µl DMTMM-modified polysaccharides for serotypes 4, 6A, 6B, 14, 18C, 19A, 19F, and 23F. Serum IgG responses to HBsAg and the Hib-PRP were quantified in a duplex fluorescent bead-based immunoassay at the same timepoints.

In May 2023, the group assignment of participants was disclosed to parents by the site study coordinator after the last child enrolled in the stage one cohort completed the scheduled visits. Except for the study statistician, the study investigators remain blinded to the group assignments of individual participants.

### Outcomes

Immunogenicity outcomes included vaccine antigen-specific IgG responses, summarised as GMCs and the proportion of participants with vaccine antigen-specific IgG concentrations meeting or exceeding established correlates of protection (i.e., 13vPCV serotype-specific IgG ≥ 0·35 μg/mL, HBsAg-IgG ≥ 10 mIU/mL, Hib-PRP-IgG ≥ 0·15 μg/mL at 6 and 7 months old and ≥ 1 μg/mL at 18 and 19 months old). Reactogenicity and parental acceptability outcomes following the 6-week, 4-month, and 6-month pertussis vaccine doses have been reported previously.^17^ Serious adverse events (SAEs) were defined according to the Australian National Health and Medical Research Council guidelines.^27^ Vaccine failure (i.e., invasive Hib disease, vaccine-serotype invasive pneumococcal disease, and hepatitis B infection) was considered an adverse event of special interest.

### Statistical analysis

The statistical analysis plan and study protocol have previously been published.^18,19^ IgG concentrations for each vaccine antigen at 6, 7, 18, and 19 months old were independently modelled using Bayesian multivariate-normal linear regression on the available log_10_ concentrations with an unstructured covariance matrix shared across both treatment groups to account for the serial correlation in the longitudinal data. The geometric mean ratio (GMR) of the mixed schedule compared to the aP-only schedule was estimated at each time point. The probability of non-inferiority was calculated using a non-inferiority margin of 2/3 on the GMR per WHO guidelines.^28^ Additionally, IgG seropositivity for each vaccine antigen was analysed via Bayesian logistic regression models with random intercepts for individuals to account for repeat measures. All models were adjusted for sex, birth order, breastfeeding status, delivery method, family history of atopic disease, and parental income as baseline covariates. Missing data were assumed to be missing at random (ignorable) under the proposed models. Analyses were conducted separately for the intent-to-treat (ITT) and per-protocol (PP) analyses sets, as prespecified in the statistical analysis plan,^19^ using R version 4·4·1 and Stan version 2·35·0 (via the CmdStanR library version 0·8·1). This trial is registered at the Australian and New Zealand Clinical Trial Registry (ACTRN12617000065392p).

## Results

Between March 7, 2018 and January 13, 2020, 153 infants were assessed in clinic for eligibility (figure 1); two were excluded (one ineligible for public funded immunisation; one received hepatitis B immunoglobulin at birth), one declined participation due to blood sampling, and 150 underwent randomisation to either the wP/aP schedule or the aP-only schedule (75 in each group). The baseline characteristics of the randomised infants were balanced across the study groups (table 1). The trial profile focuses on post-13vPCV priming and post-13vPCV boosting measurements at 6 and 18 months (earliest timepoints; figure 1); missingness patterns by assigned treatment and 18 months (earliest timepoints; figure 1); missingness patterns by assigned treatment group sets across all the scheduled phlebotomies are provided in the <u>appendix</u> (p 3).

**Figure 1.**
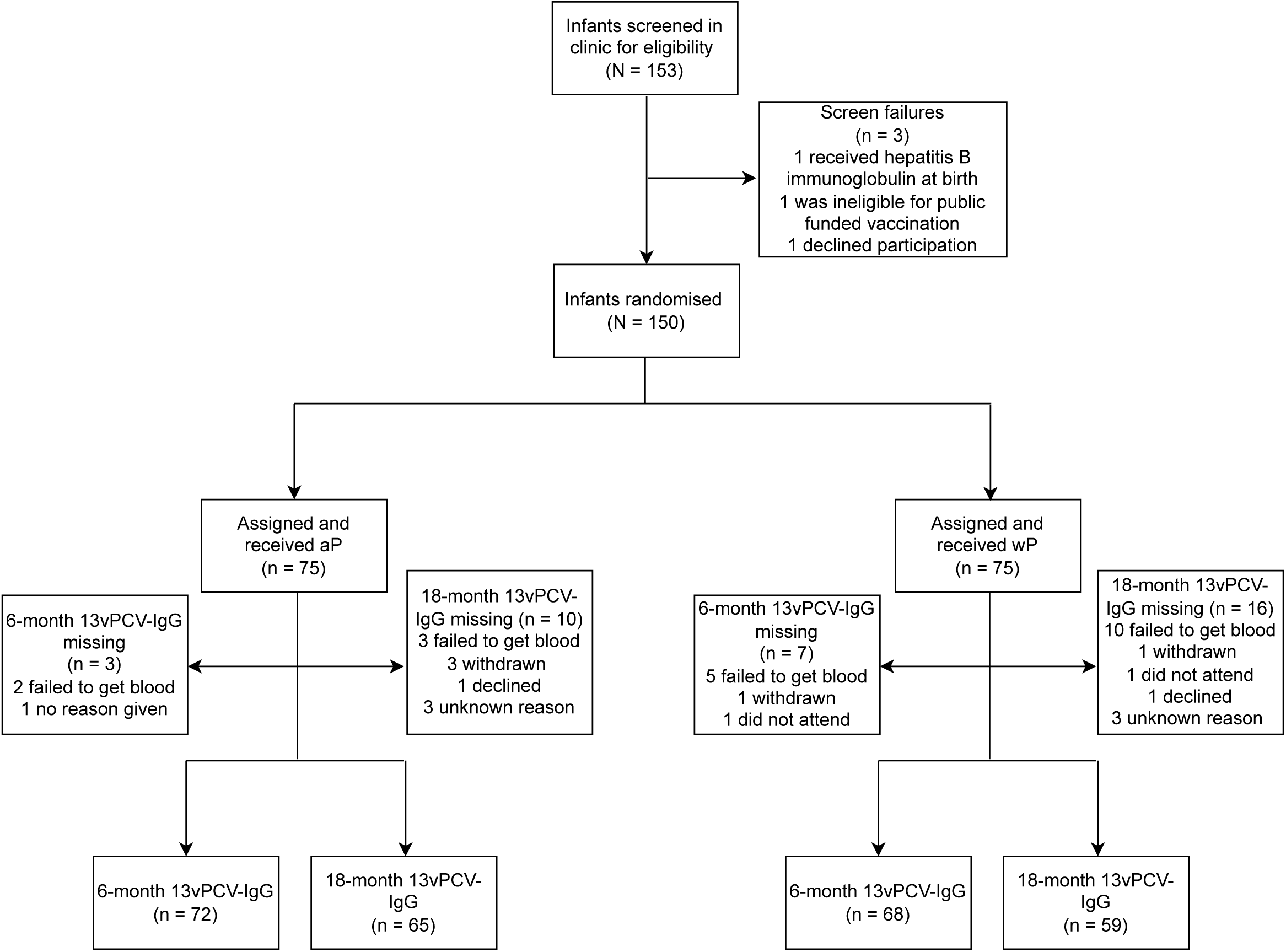
Trial profile. aP, acellular pertussis vaccine; wP, whole-cell pertussis vaccine; 13vPCV, 13-valent pneumococcal conjugate vaccine

**Table 1.** Baseline and demographic characteristics.

|  | **aP-only schedule**<br>**(n = 75)** | **Mixed wP/aP**<br>**schedule**<br>**(n = 75)** | **Overall**<br>**(n = 150)** |
| --- | --- | --- | --- |
| **Sex of child - n (%)** |  |  |  |
| Female | 36 (48) | 38 (51) | 74 (49) |
| Male | 39 (52) | 37 (49) | 76 (51) |
| **Infant's ethnicity - n (%)<sup>†</sup>** |  |  |  |
| European Caucasian | 72 (48) | 73 (49) | 145 (48) |
| Indian subcontinent | 10 (7) | 12 (8) | 22 (7) |
| Asian | 6 (4) | 11 (7) | 17 (6) |
| South American | 1 (1) | 2 (1) | 3 (1) |
| Black African | 0 (0) | 1 (1) | 1 (0) |
| Indigenous Australian | 1 (1) | 0 (0) | 1 (0) |
| **Maternal parity - n (%)** |  |  |  |
| 1 | 53 (71) | 46 (61) | 99 (66) |
| 2 | 14 (19) | 22 (29) | 36 (24) |
| 3 | 7 (9) | 4 (5) | 11 (7) |
| 4 | 1 (1) | 3 (4) | 4 (3) |
| **Maternal dTpa during this infant's pregnancy - n (%)** |  |  |  |
| 5c-dTpa | 21 (28) | 20 (27) | 41 (27) |
| 3c-dTpa | 43 (57) | 39 (52) | 82 (55) |
| Unknown | 11 (15) | 13 (17) | 24 (16) |
| None | 0 (0) | 3 (4) | 3 (2) |
| **Maternal dTpa last 5 years, prior to this infant's pregnancy - n (%)** |  |  |  |
| 5c-dTpa | 7 (9) | 6 (8) | 13 (9) |
| 3c-dTpa | 6 (8) | 9 (12) | 15 (10) |
| 3c-dTpa-IPV | 1 (1) | 0 (0) | 1 (1) |
| Unknown | 22 (29) | 25 (33) | 47 (31) |
| None | 39 (52) | 35 (47) | 74 (49) |

<sup>†</sup> Multiple ethnicities may apply, so percentages may not sum to 100. aP: acellular pertussis vaccine. wP: whole-cell pertussis vaccine as a first dose. 5c-dTpa: 5-component diphtheria-tetanus-acellular pertussis combination vaccine (reduced antigen formulation; includes pertussis toxoid, filamentous haemagglutinin, pertactin, and fimbriae types 2 and 3); 3c-dTpa: 3-component diphtheria-tetanus-acellular pertussis combination vaccine (reduced antigen formulation; includes pertussis toxoid, filamentous haemagglutinin, and pertactin); IPV: inactivated poliovirus vaccine.

At 6 months old, the lowest pneumococcal serotype-specific IgG GMCs in both study groups were for serotypes 3, 6B, 19A, and 23F (ITT analysis set, table 2 and <u>appendix</u> p 4; PP analysis set, <u>appendix</u> pp 10–11). The probability of the GMR exceeding 2/3 (evaluating non-inferiority) in the wP/aP group compared to the aP-only group was > 0.99 for all 13 vaccine serotypes in both ITT (figure 2; <u>appendix</u> pp. 13–14) and PP analysis sets (<u>appendix</u> pp 15–17). The probability of the GMR exceeding 1 (evaluating superiority of wP/aP) ranged from 0.88 to > 0.99 in the ITT (figure 2; <u>appendix</u> pp. 13–14) and in the PP analysis set (<u>appendix</u> pp 15–17) for all serotypes. For each of the 13 vaccine serotypes, the probability of a seroprotection difference > 0 (evaluating superiority of wP/aP) in the wP/aP group compared to the aP-only group ranged from 0·80 to > 0·99 in the ITT analysis set (figure 3 and <u>appendix</u> pp. 18–19) and from 0·71 to > 0·99 in the PP analysis set (<u>appendix</u> pp. 20– 22). Similar findings were observed at 7 months old (3 months after 13vPCV priming); these data are presented in table 2, figures 2 and 3, and in the <u>appendix</u> (p 4, pp 10–11, and 18– 22).

**Figure 2.**
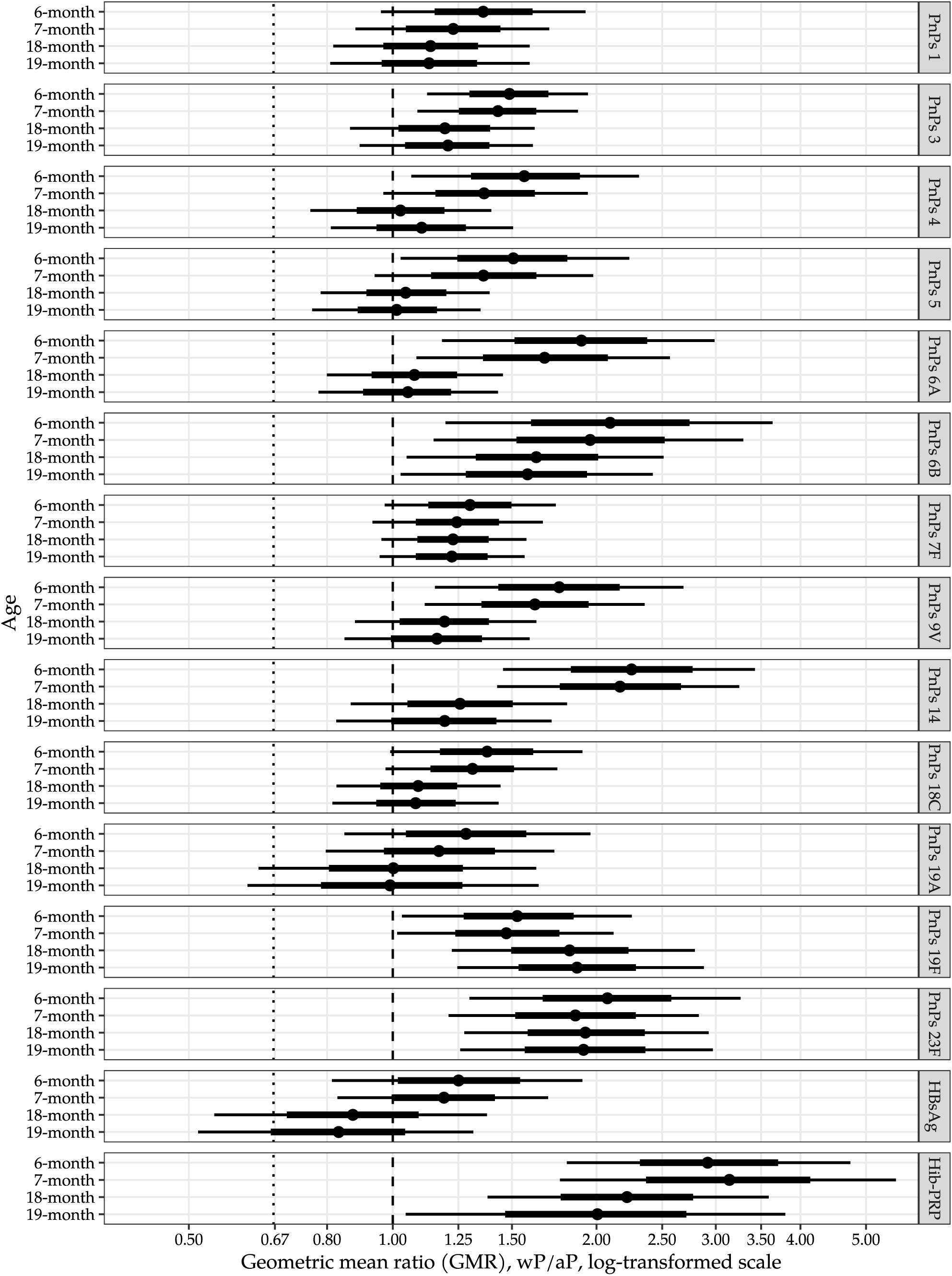
IgG geometric mean ratio (GMR, wP/aP vs aP-only) by vaccination age and antigen/serotype (intention-to-treat). Points - median, rectangles - 80% credible interval (CrI), lines - 95% CrI. *aP, acellular pertussis vaccine; wP, whole-cell pertussis vaccine; PnPs, pneumococcal polysaccharide; HBsAg, hepatitis B surface antigen; Hib-PRP, Haemophilus influenzae type b capsular polysaccharide polyribosylribitol phosphate*.

**Figure 3.**
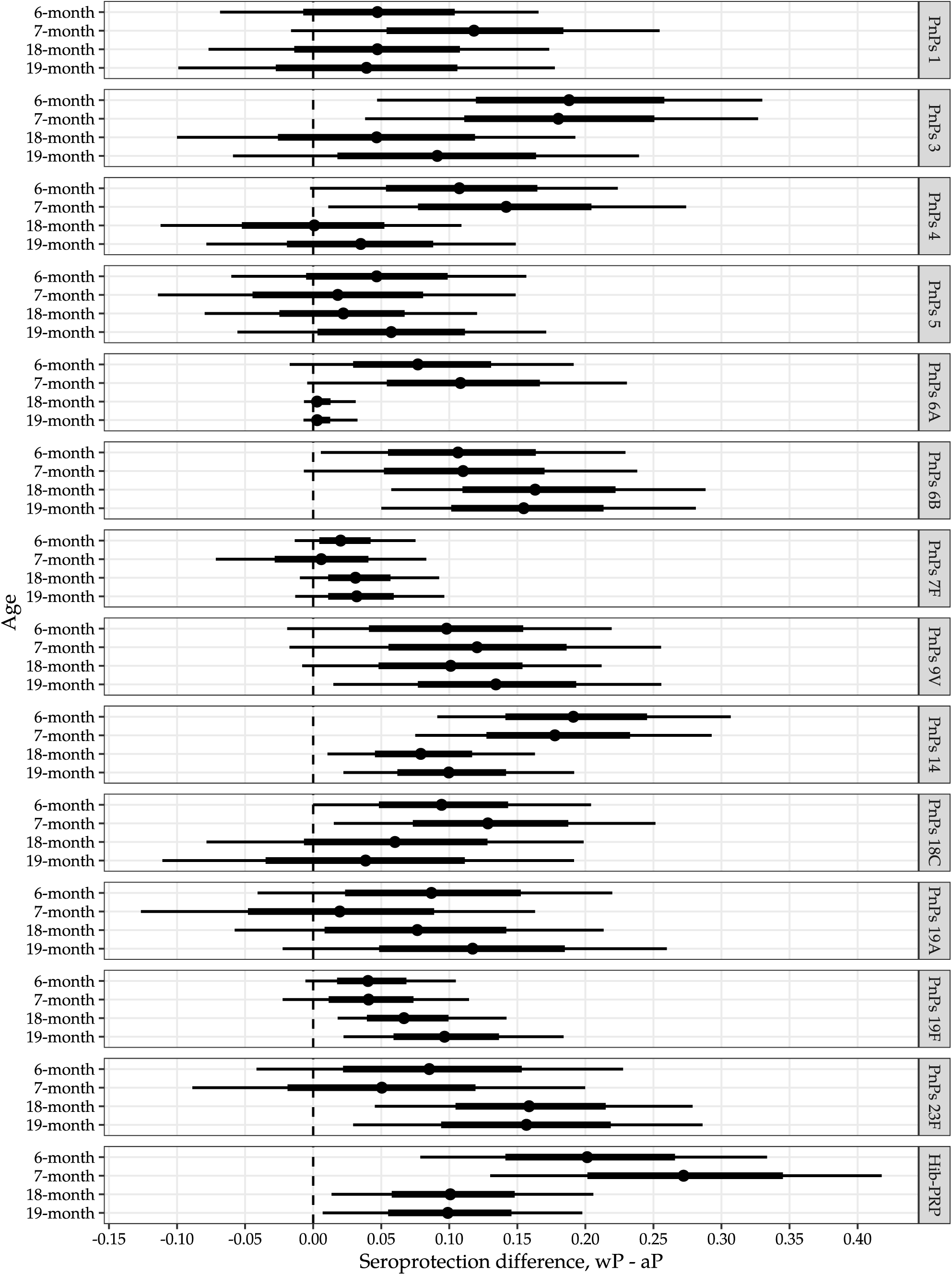
Standardised probability difference (wP/aP - aP-only) for IgG seroprotection by serotype/antigen, age, and assigned treatment (intention-to-treat). Points - median, rectangles - 80% credible interval (CrI), lines - 95% CrI. *aP, acellular pertussis vaccine; wP, whole-cell pertussis vaccine; PnPs, pneumococcal polysaccharide; Hib-PRP, Haemophilus influenzae type b capsular polysaccharide polyribosylribitol phosphate*

**Table 2.** Descriptive statistics of IgG concentrations by antigen type, age and assigned treatment group.

|  | aP-only (n = 75) |  |  |  |  |  | wP/aP (n = 75) |  |  |  |  |  |
| --- | --- | --- | --- | --- | --- | --- | --- | --- | --- | --- | --- | --- |
|  | n | GMC<sup>1</sup> | GSD<sup>2</sup> | Q1–Q3<sup>3</sup> | Min–Max | S+<sup>4</sup> | n | GMC<sup>1</sup> | GSD<sup>2</sup> | Q1–Q3<sup>3</sup> | Min–Max | S+<sup>4</sup> |
| **PnPs 1** |  |  |  |  |  |  |  |  |  |  |  |  |
| 6-month | 72 | 0.63 | 3.02 | 0.29–1.56 | 0.04–10.5 | 68% | 68 | 0.86 | 2.83 | 0.35–1.93 | 0.16–8.85 | 74% |
| 7-month | 69 | 0.51 | 2.87 | 0.24–1.11 | 0.04–7.26 | 55% | 67 | 0.68 | 2.74 | 0.31–1.50 | 0.10–4.87 | 72% |
| 18-month | 65 | 0.55 | 2.86 | 0.25–0.95 | 0.07–7.37 | 68% | 59 | 0.69 | 2.66 | 0.36–1.25 | 0.09–8.23 | 76% |
| 19-month | 64 | 0.48 | 2.89 | 0.22–0.89 | 0.06–8.77 | 61% | 54 | 0.65 | 2.54 | 0.31–1.16 | 0.11–5.14 | 72% |
| **PnPs 3** |  |  |  |  |  |  |  |  |  |  |  |  |
| 6-month | 72 | 0.28 | 2.39 | 0.19–0.47 | 0.02–1.89 | 39% | 68 | 0.43 | 2.22 | 0.29–0.70 | 0.05–2.06 | 63% |
| 7-month | 69 | 0.22 | 2.32 | 0.12–0.33 | 0.02–1.16 | 23% | 67 | 0.33 | 2.30 | 0.20–0.52 | 0.05–2.58 | 45% |
| 18-month | 65 | 0.25 | 2.67 | 0.13–0.51 | 0.03–2.35 | 35% | 59 | 0.32 | 2.42 | 0.18–0.48 | 0.06–4.52 | 41% |
| 19-month | 64 | 0.22 | 2.60 | 0.12–0.40 | 0.03–3.33 | 30% | 54 | 0.30 | 2.19 | 0.15–0.43 | 0.09–2.25 | 41% |
| **PnPs 4** |  |  |  |  |  |  |  |  |  |  |  |  |
| 6-month | 72 | 0.66 | 3.60 | 0.29–1.64 | 0.02–10.5 | 69% | 68 | 1.03 | 3.13 | 0.54–2.09 | 0.04–7.64 | 85% |
| 7-month | 69 | 0.54 | 3.26 | 0.25–1.21 | 0.02–9.18 | 62% | 67 | 0.82 | 2.68 | 0.44–1.54 | 0.04–4.72 | 81% |
| 18-month | 65 | 0.74 | 2.42 | 0.45–1.46 | 0.09–8.31 | 83% | 59 | 0.84 | 2.46 | 0.50–1.47 | 0.09–14.0 | 83% |
| 19-month | 64 | 0.61 | 2.45 | 0.40–1.22 | 0.06–10.6 | 80% | 54 | 0.80 | 2.43 | 0.50–1.39 | 0.14–13.2 | 85% |
| **PnPs 5** |  |  |  |  |  |  |  |  |  |  |  |  |
| 6-month | 72 | 0.53 | 3.69 | 0.17–1.40 | 0.02–8.19 | 72% | 68 | 0.82 | 2.96 | 0.44–1.77 | 0.03–5.37 | 81% |
| 7-month | 69 | 0.50 | 3.35 | 0.22–1.20 | 0.03–4.89 | 70% | 67 | 0.72 | 2.81 | 0.34–1.50 | 0.03–4.75 | 73% |
| 18-month | 65 | 0.72 | 2.28 | 0.44–1.27 | 0.15–5.39 | 83% | 59 | 0.83 | 2.25 | 0.55–1.31 | 0.09–9.46 | 88% |
| 19-month | 64 | 0.67 | 2.23 | 0.38–1.08 | 0.14–5.16 | 77% | 54 | 0.78 | 2.20 | 0.50–1.27 | 0.11–6.22 | 89% |
| **PnPs 6A** |  |  |  |  |  |  |  |  |  |  |  |  |
| 6-month | 72 | 0.87 | 4.96 | 0.31–2.83 | 0.01–30.5 | 72% | 68 | 1.60 | 3.87 | 0.81–4.22 | 0.02–14.4 | 85% |
| 7-month | 69 | 0.75 | 4.41 | 0.25–2.21 | 0.03–18.1 | 70% | 67 | 1.39 | 3.38 | 0.75–3.45 | 0.03–9.41 | 88% |
| 18-month | 65 | 2.42 | 2.51 | 1.24–4.57 | 0.43–13.0 | 100% | 59 | 2.78 | 2.35 | 1.55–4.14 | 0.55–45.5 | 100% |
| 19-month | 64 | 2.26 | 2.60 | 1.11–4.26 | 0.38–16.4 | 100% | 54 | 2.57 | 2.36 | 1.48–3.40 | 0.42–32.2 | 100% |
| **PnPs 6B** |  |  |  |  |  |  |  |  |  |  |  |  |
| 6-month | 72 | 0.06 | 5.60 | 0.02–0.21 | 0.00–5.49 | 14% | 68 | 0.14 | 5.71 | 0.03–0.40 | 0.01–14.2 | 29% |
| 7-month | 69 | 0.07 | 4.95 | 0.02–0.25 | 0.00–4.25 | 17% | 67 | 0.14 | 5.27 | 0.04–0.46 | 0.01–8.96 | 31% |
| 18-month | 65 | 0.85 | 4.01 | 0.30–2.41 | 0.03–8.17 | 72% | 59 | 1.54 | 3.08 | 0.69–3.13 | 0.16–100 | 93% |
| 19-month | 64 | 0.81 | 4.07 | 0.29–2.62 | 0.03–15.5 | 73% | 54 | 1.44 | 2.93 | 0.68–2.80 | 0.18–62.4 | 94% |
| **PnPs 7F** |  |  |  |  |  |  |  |  |  |  |  |  |
| 6-month | 72 | 1.31 | 2.60 | 0.84–2.60 | 0.06–10.6 | 94% | 68 | 1.79 | 2.38 | 1.06–3.26 | 0.07–8.35 | 99% |
| 7-month | 69 | 1.12 | 2.61 | 0.73–2.23 | 0.04–6.85 | 91% | 67 | 1.47 | 2.36 | 0.84–2.94 | 0.07–6.32 | 94% |
| 18-month | 65 | 1.36 | 2.09 | 0.86–2.24 | 0.25–6.53 | 94% | 59 | 1.76 | 1.93 | 1.12–2.49 | 0.39–18.7 | 100% |
| 19-month | 64 | 1.34 | 2.13 | 0.94–2.08 | 0.23–8.76 | 94% | 54 | 1.79 | 1.86 | 1.21–2.51 | 0.47–13.5 | 100% |
| **PnPs 9V** |  |  |  |  |  |  |  |  |  |  |  |  |
| 6-month | 72 | 0.49 | 4.39 | 0.26–1.50 | 0.02–14.8 | 68% | 68 | 0.87 | 3.14 | 0.45–1.96 | 0.04–6.62 | 79% |
| 7-month | 69 | 0.43 | 3.56 | 0.20–1.07 | 0.02–6.72 | 59% | 67 | 0.73 | 2.74 | 0.35–1.52 | 0.04–5.18 | 75% |
| 18-month | 65 | 0.71 | 2.67 | 0.35–1.59 | 0.11–6.15 | 75% | 59 | 0.90 | 2.31 | 0.48–1.56 | 0.14–8.86 | 85% |
| 19-month | 64 | 0.64 | 2.72 | 0.29–1.22 | 0.11–9.05 | 69% | 54 | 0.83 | 2.38 | 0.40–1.41 | 0.13–5.66 | 83% |
| **PnPs 14** |  |  |  |  |  |  |  |  |  |  |  |  |
| 6-month | 72 | 1.06 | 4.21 | 0.26–3.71 | 0.07–15.0 | 69% | 68 | 2.38 | 3.23 | 1.20–4.76 | 0.05–17.1 | 96% |
| 7-month | 69 | 1.02 | 4.20 | 0.39–2.86 | 0.04–29.3 | 75% | 67 | 2.11 | 2.94 | 1.23–4.68 | 0.06–15.8 | 94% |
| 18-month | 65 | 1.51 | 3.14 | 0.62–3.39 | 0.09–15.9 | 91% | 59 | 1.89 | 2.41 | 1.09–3.61 | 0.05–14.8 | 97% |
| 19-month | 64 | 1.33 | 3.17 | 0.57–2.73 | 0.09–17.6 | 88% | 54 | 1.65 | 2.49 | 1.14–3.12 | 0.04–10.4 | 96% |
| **PnPs 18C** |  |  |  |  |  |  |  |  |  |  |  |  |
| 6-month | 72 | 0.76 | 2.68 | 0.44–1.53 | 0.06–7.64 | 78% | 68 | 1.03 | 2.63 | 0.60–2.04 | 0.05–6.08 | 90% |
| 7-month | 69 | 0.58 | 2.43 | 0.31–1.14 | 0.08–5.22 | 70% | 67 | 0.82 | 2.36 | 0.48–1.49 | 0.06–4.97 | 87% |
| 18-month | 65 | 0.49 | 2.28 | 0.28–0.85 | 0.09–4.06 | 65% | 59 | 0.58 | 2.20 | 0.36–1.04 | 0.11–9.65 | 76% |
| 19-month | 64 | 0.45 | 2.32 | 0.24–0.80 | 0.09–4.52 | 58% | 54 | 0.55 | 2.27 | 0.32–0.98 | 0.12–9.61 | 70% |
| **PnPs 19A** |  |  |  |  |  |  |  |  |  |  |  |  |
| 6-month | 72 | 0.36 | 3.45 | 0.12–0.90 | 0.04–5.78 | 54% | 68 | 0.47 | 3.71 | 0.21–1.15 | 0.01–6.13 | 66% |
| 7-month | 69 | 0.27 | 3.08 | 0.09–0.63 | 0.04–4.31 | 46% | 67 | 0.33 | 3.35 | 0.15–0.79 | 0.01–4.59 | 51% |
| 18-month | 65 | 0.67 | 4.21 | 0.24–1.42 | 0.07–164 | 65% | 59 | 0.75 | 3.43 | 0.34–1.33 | 0.06–44.0 | 75% |
| 19-month | 64 | 0.60 | 4.61 | 0.21–1.19 | 0.07–158 | 58% | 54 | 0.73 | 3.52 | 0.38–1.40 | 0.05–37.7 | 76% |
| **PnPs 19F** |  |  |  |  |  |  |  |  |  |  |  |  |
| 6-month | 72 | 2.20 | 3.65 | 1.15–5.64 | 0.02–27.2 | 93% | 68 | 3.33 | 3.42 | 1.96–7.16 | 0.01–36.1 | 97% |
| 7-month | 69 | 1.55 | 3.53 | 0.91–3.84 | 0.01–13.9 | 91% | 67 | 2.49 | 3.11 | 1.50–4.30 | 0.01–30.5 | 97% |
| 18-month | 65 | 1.45 | 3.72 | 0.58–3.41 | 0.07–112 | 91% | 59 | 2.81 | 3.03 | 1.47–4.77 | 0.52–57.7 | 100% |
| 19-month | 64 | 1.24 | 3.61 | 0.53–3.30 | 0.08–101 | 86% | 54 | 2.71 | 3.24 | 1.49–4.91 | 0.30–145 | 98% |
| **PnPs 23F** |  |  |  |  |  |  |  |  |  |  |  |  |
| 6-month | 72 | 0.24 | 4.60 | 0.11–0.59 | 0.00–4.91 | 44% | 68 | 0.49 | 4.03 | 0.19–1.66 | 0.01–6.58 | 56% |
| 7-month | 69 | 0.19 | 4.17 | 0.09–0.49 | 0.00–2.83 | 38% | 67 | 0.39 | 3.64 | 0.17–1.04 | 0.01–4.97 | 51% |
| 18-month | 65 | 0.63 | 3.34 | 0.33–1.61 | 0.04–5.73 | 71% | 59 | 1.35 | 3.68 | 0.55–3.07 | 0.14–72.3 | 86% |
| 19-month | 64 | 0.62 | 3.41 | 0.29–1.48 | 0.05–10.8 | 64% | 54 | 1.37 | 3.57 | 0.56–3.32 | 0.17–44.1 | 83% |
| **HBsAg** |  |  |  |  |  |  |  |  |  |  |  |  |
| 6-month | 72 | 3.81 | 3.78 | 1.91–9.00 | 0.04–33.7 | 100% | 68 | 4.80 | 3.66 | 2.32–12.2 | 0.06–51.4 | 100% |
| 7-month | 69 | 7.53 | 2.90 | 3.97–14.8 | 0.24–42.3 | 100% | 67 | 8.66 | 3.08 | 3.82–20.5 | 0.53–51.8 | 100% |
| 18-month | 65 | 0.52 | 4.14 | 0.21–1.49 | 0.02–7.59 | 100% | 59 | 0.49 | 4.04 | 0.22–1.34 | 0.01–14.2 | 100% |
| 19-month | 64 | 0.46 | 4.33 | 0.20–1.27 | 0.02–7.06 | 100% | 54 | 0.40 | 3.81 | 0.21–1.13 | 0.02–8.35 | 100% |
| **Hib-PRP** |  |  |  |  |  |  |  |  |  |  |  |  |
| 6-month | 72 | 0.04 | 3.71 | 0.02–0.06 | 0.00–1.98 | 12% | 68 | 0.11 | 4.85 | 0.03–0.28 | 0.01–9.03 | 38% |
| 7-month | 69 | 0.20 | 5.54 | 0.07–0.67 | 0.00–8.28 | 54% | 67 | 0.64 | 5.14 | 0.27–2.43 | 0.02–28.3 | 82% |
| 18-month | 65 | 0.09 | 3.11 | 0.04–0.20 | 0.01–1.62 | 3% | 59 | 0.22 | 4.69 | 0.07–0.48 | 0.01–21.4 | 15% |
| 19-month | 64 | 10.18 | 5.71 | 3.57–36.5 | 0.06–153 | 84% | 54 | 23.25 | 6.26 | 8.98–65.7 | 0.13–984 | 93% |

*aP*, acellular pertussis vaccine; *wP*, whole-cell pertussis vaccine; <sup>1</sup>*GMC*, geometric mean concentration; <sup>2</sup>*GSD*, geometric standard deviation, <sup>3</sup>*Q1 – 25<sup>th</sup> sample percentile*, *Q3 – 75<sup>th</sup> sample percentile*; *Min–Max*, minimum-maximum; <sup>4</sup>*S+*, seroprotective at specified level; *PnPs*, pneumococcal polysaccharide; *Hib-PRP*, *Hib-PRP*, *Haemophilus influenzae* type *b* capsular polysaccharide polyribosylribitol phosphate; *HBsAg*, hepatitis B surface antigen

At 18 months old (6 months post-13vPCV booster), serotypes 1, 3, 18C, and 19A had the lowest pneumococcal serotype-specific IgG GMCs across both groups (ITT analysis set, table 2 and <u>appendix</u> p 4; PP analysis set, <u>appendix</u> pp 10–11). The probability of the GMR exceeding 2/3 in the wP/aP group compared to the aP-only group ranged from 0·96 to > 0·99 for all 13 vaccine serotypes in the ITT analysis set (figure 2; <u>appendix</u> pp. 13–14) and from 0·94 to > 0·99 in the PP analysis set (<u>appendix</u> pp. 15–16). Additionally, the probability of the GMR exceeding 1 ranged from 0·50 to > 0·99 in the ITT (figure 2; <u>appendix</u> pp. 13– 14), with the lowest for serotype 19A, and from 0·14 to > 0·99 in the PP analysis set (<u>appendix</u> pp. 15–17), with the lowest for serotype 4. For each of the 13 vaccine serotypes, the probability of a seroprotection difference > than 0 in the wP/aP group compared to the aP-only group ranged from 0·51 to > 0·99 in the ITT analysis set (figure 3; <u>appendix</u> pp. 18– 19) and from 0·42 to > 0·99 in the PP analysis set (<u>appendix</u> pp 20–22), with the lowest for serotypes 3, 4, and 5. Similar findings were observed at 19 months old (7 months post-13vPCV booster); these data are presented in table 2, figures 2 and 3, and in the <u>appendix</u> (p 4, pp 10–11, 18–22).

In the ITT analysis set, both study groups exhibited low GMCs of anti-Hib-PRP immediately before the third dose of the Hib-PRP vaccine at age 6 months (wP/aP group = 0·11 µg/mL versus aP-only group = 0·04 µg/mL). A minority had anti-PRP IgG ≥ 0·15 μg/mL (wP/aP, n = 26/68 [38%]; aP-only, n = 9/72 [12%], table 2; <u>appendix</u> p 5). By 7 months old (4 weeks post Hib-PRP priming), IgG GMCs and seroprotection rates were higher in the wP/aP group (0·64 µg/mL; n = 55/67 [82%]) compared to the aP-only group (0·20 µg/mL; n = 37/69 [54%]; table 2; <u>appendix</u> p 5). GMCs for Hib-PRP IgG were both non-inferior (GMR > 2/3) and superior (GMR > 1) with a probability > 0·99 in the wP/aP group compared to the aP-only group at both ages (figure 2; <u>appendix</u> pp 13–14). The probability of a seroprotection difference > than 0 in the wP/aP group compared to the aP-only group was also > 0·99 at both 6 and 7 months (figure 3; <u>appendix</u> pp 18–19). Similar results were observed in the PP analysis set (<u>appendix</u> pp 15–16; pp 20–21).

Anti-Hib-PRP IgG GMCs declined between 7 months and immediately before the Hib-PRP booster dose at 18 months, from 0·64 to 0·22 µg/mL in the wP/aP group and from 0·20 to 0·09 µg/mL in the aP-only group (ITT analysis set, table 2). At 18 months, few toddlers had Hib-PRP IgG levels ≥ 1·00 μg/mL (wP/aP, n = 9/59 [15%] versus aP-only, n = 2/65 [3%]). By 19 months (4 weeks post-Hib-PRP boosting), IgG GMCs and seroprotection rates were higher in the wP/aP group (23·25 µg/mL; n = 50/54 [93%]) compared to the aP-only group (10·18 µg/mL; n = 54/64 [84%]) (table 2). GMCs for Hib-PRP were both non-inferior (GMR > 2/3) and superior (GMR > 1) with a probability > 0·99 in the wP/aP group versus the aP-only group (figure 2; <u>appendix</u> pp.13–14). The probability of seroprotection difference > 0 in the wP/aP group compared to the aP-only group was 0.99 and 0.98 at 18 and 19 months, respectively. Similar results were observed in the PP analysis set (<u>appendix</u> pp.12, 15–17, 20–22). After Hib-PRP boosting, 50 of 54 toddlers (93%) in the wP/aP group and 62 of 64 (97%) in the aP-only group achieved a fourfold seroconversion in Hib-PRP IgG. The probability of a difference > 0 in the wP/aP group compared to the aP-only group was 0·21 and 0·55 in the ITT and PP analysis sets (<u>appendix</u> p 23–24), respectively.

At 6 months (pre-third dose of hepatitis B vaccine) and 7 months old (4 weeks post-hepatitis B priming), all infants across the study groups had HBsAg IgG concentrations above the specified seroprotective threshold in both ITT (table 2; appendix p 5) and PP analyses sets (<u>appendix</u> p 12). GMCs for HBsAg IgG were non-inferior (GMR > 2/3) with at least 95% probability in the wP/aP group compared to the aP-only group at both ages (<u>appendix</u> pp. 13–17).

No episodes of vaccine-serotype invasive pneumococcal disease, invasive Hib disease, or hepatitis B infection were ascertained during the follow-up period (∼ 18 months). Additional SAEs and adverse events of special interest (breakthrough pertussis infections and hypotonic hyporesponsive episodes) occurring within the first 6 months of follow-up were described previously;^17^ SAEs following the 18-month vaccine doses and unsolicited adverse events by treatment group will be reported at the end of the trial.

## Discussion

Here we report for the first time, the long-term immunomodulatory effect of a mixed wP/aP schedule on co-administered vaccine responses up to 19 months old. In this randomised study, compared to an aP-only schedule, a mixed wP/aP schedule resulted in non-inferior IgG responses to co-administered pneumococcal vaccine serotypes and Hib-PRP at ages 6, 7, 18, and 19 months, and to HBsAg at 6 and 7 months old, all with high probability. In many instances, antibody responses (IgG GMR) were superior in the mixed wP/aP schedule group (i.e., 6B, 19F, and 23F at all the assessed time points). Consistent with earlier reports,^10,29^ we hypothesise that the innate immune activation caused by the pathogen associated molecular patterns in the whole pertussis bacteria present in the wP vaccine, along with the subsequent activation of antigen-presenting cells and pro-inflammatory cytokines, provides stronger signals to stimulate CD4^+^ T cells. Further these stimuli support the induction and maturation of polysaccharide-specific memory B cells and plasma cell generation, as well as memory B cell and plasma cell responses with broader isotypic profiles, providing robust and more durable protective immunity. Live attenuated vaccines, such as those for measles, mumps, rubella, and BCG, have also been suggested to boost immune responses to unrelated pathogens.^30^ The proposed mechanisms for BCG’s non-specific effects include epigenetic reprogramming of monocytes and a metabolic shift in innate immune cells in both animal experiments and ex vivo studies in humans.^31^ In the future, we will assess additional stored sera and cryopreserved peripheral blood mononuclear cells from this cohort using functional opsonophagocytic assays, global T-cell and memory B-cell phenotyping to determine the functional impact and cellular drivers of the differences observed here.

The strengths of our trial lie in its rich data collection allowing us to measure individual-level antibody trajectories across four time points while randomisation minimised potential bias from imbalance in baseline demographics between the two schedules. This approach enabled detailed insights into the immunological outcomes of the different vaccination schedules. The study also derived comprehensive reactogenicity and parental acceptability data (which has been reported previously) that demonstrates the wP/aP strategy (using a WHO-prequalified vaccine formulation) is well-tolerated and accepted, albeit more reactogenic.^17^ Our previous meta-analysis found that the risk difference for SAEs in receiving a first dose of wP versus aP is likely small, ranging from three fewer to two more events per 1,000 children.^32^ As described previously, none of the seven SAE reported within the first six months of follow-up among five infants in the OPTIMUM stage one cohort were assessed to be related to the study vaccines.^17^

Our study has three main limitations. First, we did not collect baseline (pre-vaccination) 6-week samples to measure passively transferred maternal antibody levels which are known to also influence infant vaccine immunogenicity.^33,34^ Exploratory analyses on this cohort, including history of maternal dTpa as well as data from simultaneous observational studies involving infants primed with the standard aP-only schedule may provide further insights on the role of immune interference by maternal antibodies in this setting. Second, the phlebotomy schedule was driven by parsimony and the priority to assess the immunogenicity of the pertussis priming schedule. We were therefore not able to measure peak immune responses to PCV serotypes which are likely to have occurred approximately one month after the 4-month and 12-month 13vPCV doses. Instead, we could only examine the impact of mixed wP/aP priming at two- and three-months post-primary 13vPCV, and at six months post-boosting, respectively. Third, it was beyond the scope of this study to assess the implications of differential immune responses on risk of disease; this will require very large population-level analyses. Fourth, while the adjuvanticity of IPV as part of DTaP/dTpa formulations has been reported previously in Australian and European cohorts,^9,35,36^ we were unable to further explore the proposed immunostimulant effects of IPV co-vaccination on Hib-PRP and 13vPCV responses.

An aggregate correlate of protection of 0.35 µg/ml is established for protection against invasive pneumococcal disease;^37^ however higher IgG concentrations appear to be required for protection against individual serotypes and specific populations.^38^ In the 3 + 0 and 2 + 1 PCV schedule era, serotypes 3 and 19A have been the major etiological agents of breakthrough invasive pneumococcal disease in Australia despite high vaccine coverage.^39–41^ Here we find that regardless of pertussis vaccine primary series strategy, serotype 3 was associated with the lowest GMCs when measured during the second year of life. While the GMC and proportion meeting the seroprotective threshold for these serotypes were numerically higher for the mixed wP/aP group at each timepoint, the data is inconclusive for an improved immunogenic response for these specific serotypes. Finally, the multiplexed immunoassays used here have been developed for research purposes and the established seroprotective thresholds may not perform similarly across technologies; the data therefore should be used to interpret immune kinetics only.

We conclude that the mixed wP/aP effectively boosts IgG responses to co-administered vaccine antigens and enhances the booster effect of polysaccharide vaccine antigens given in the second year of life. While the implications for vaccine-preventable disease are uncertain, this evidence supports countries with stable wP-based pertussis and 13vPCV programs to follow WHO recommendations for the continued use of wP schedules. These finding are also anticipated to inform 13vPCV dose-sparing policies (e.g., from 3 + 0 to 2 +1 PCV schedules) globally.

## Data Availability

Data from the study will be available at the completion of follow-up, analysis and reporting of OPTIMUM stage two; no end date. Deidentified, individual participant data and a data dictionary defining each field in the set, will be made available to researchers who provide a methodologically sound proposal to the University of Sydney, Australia (Ms Nicole Wong, Clinical Trial Support Lead; Department: Pro Vice-Chancellor, Research), subject to a signed data access agreement and any necessary ethics approvals (see ANZCTR trial registration). The study protocol and statistical analysis plan are published, other study related documentation is available on request.

https://osf.io/qxkhe/?view_only=7bb25f8bd0044238b3e464ca2547d6bc

## Contributors

Conceptualisation: MJE, JAM, DEC, MG, CSW, NC, PGH, TS, PCR. Data curation: GPC, JT. Formal analysis: SMc, JT. Funding acquisition: MJE, JAM, KPP, DEC, MG, CSW, NC, PBM, PGH, TS, PCR. Investigation: GPC, DEC, MoS, UW. Methodology: SMc, JT, JAM, NC, TS, PCR. Project administration: MJE. Software: JT. Supervision: JT, MJE, TS, PCR. Validation: SMc, JT, MJ. Visualisation: JT. Data interpretation: GPC, SMc, JT, MJE, MJ, KPP, NW, CSW, UW, PBM, TS, PCR. Writing – original draft: GPC. Writing – review & editing: All authors. JT and MJ have directly accessed and verified the underlying data reported in the manuscript.

## Acknowledgments

We acknowledge Aboriginal and Torres Strait Islander People as the Traditional Custodians of the land and waters of Australia. We also acknowledge the Nyoongar Wadjuk, Yawuru, Kariyarra, and Kaurna Elders, their people and their land upon which The Kids Research Institute Australia is located, and where stage one of the OPTIMUM study was conducted. We seek their wisdom in our work to improve the health and development of all children.

We thank the parents and infants who took part in stage one of this trial for their commitment and continuous support. We acknowledge all members of the Vaccine Trials Group at The Kids Research Institute Australia, who were involved in the conduct of stage one of the OPTIMUM study. We would also like to express our gratitude to Kylie Rogers (Data Manager) and Prof Kathryn M. Edwards, MD (Professor of Pediatrics Emerita, Vanderbilt University School of Medicine, USA) for their valuable contributions. Additionally, we thank the independent Data Safety Monitoring Committee (Chair – Prof Keith Grimwood, Emeritus Prof Don Roberton (former Chair), Prof Allen Cheng, Prof Nigel Crawford, Prof David Isaacs, Associate Prof Stephen Lambert, Prof Helen Marshall, Prof Emma McBryde, Dr Sam Mehr, and Associate Prof John Reynolds) for their detailed oversight. Our appreciation extends to Catherine Hughes (AM), consumer representative in the OPTIMUM study Steering Committee for her insightful input in the design and conduct of this study.

This is an investigator-initiated study supported by grants from the NHMRC (GNT1158722) awarded to TS, DEC, MG, JAM, PCR, NW, and KPP, and the Telethon New Children’s Hospital Research Fund 2012 awarded to TS, PCR, and PGH. GPC was supported by PhD scholarships from the Endeavour Leadership Program (Australian Department of Education and Training), the Forrest Research Foundation, and the Wesfarmers Centre of Vaccines and Infectious Diseases during the conduct of the study. NC is supported by a NHMRC investigator grant (GNT1197117). KPP is supported by a NHMRC fellowship (GNT2008911), and a Melbourne Children’s Clinician-Scientist Fellowship. Research at MCRI is supported by the Victorian Government’s Operational Infrastructure Support Program. TS is supported by a Medical Research Future Fund Investigator Grant (MRF119515).

